# Socio-economic determinants of early antenatal care visit among pregnant women in Zambia (2007-2019): Evidence from the Zambia Demographic and Health Surveys

**DOI:** 10.1101/2024.03.08.24303972

**Authors:** Wingston Felix Ng’ambi, Cosmas Zyambo, Alice Ngoma Hazemba, Adamson Sinjani Muula, Dominic Nkhoma, Twaambo Hamonga, Angel Mwiche

**Affiliations:** Kamuzu University of Health Sciences, Department of Health Systems and Policy, Health Economics and Policy Unit, Lilongwe, Malawi; University of Zambia, Department of Public Health and Family Medicine, Lusaka, Zambia; Kamuzu University of Health Sciences, School of Public Health, Blantyre, Malawi; Ministry of Health, Reproductive Health Directorate, Lusaka, Zambia

**Keywords:** SSA, Zambia, ANC, ANC4+

## Abstract

**INTRODUCTION:** The timing of antenatal care (ANC) attendance may affect outcomes for mother and child health. Using the Zambia Demographic and Health Survey (ZDHS), we describe the adoption of at least four early ANC (ANC4+) visits and early uptake of ANC among women of reproductive age in Zambia between 2007 and 2019.

**METHODS:** We made use of ZDHS data gathered between 2007 and 2019. In this investigation, all women between the ages of 15 and 49 were taken into account. Early ANC4+ was the desired result, which was defined as having at least four ANC visits with the first ANC visit occurring during the first four months of pregnancy. In Stata v17, weighted univariate, bivariate, and multivariate logistic regression analyses were performed.

**RESULTS:** A total of 11633 (56%) of the 20661 women enrolled in our study had received early initiation of ANC4+. We saw an increase in the proportion of women who started ANC4+ early, from 55% in 2007 to 63% in 2018/19. There was a decreasing trend in the odds of early ANC4+ initiation with parity, but an increasing trend in the odds of early ANC4+ initiation with a higher level of education. Being a member of a wealthier household was associated with a lower risk of ANC4+ (OR= 0.81, 95%CI: 0.66-0.99, P=0.03). Twenty-seven percent of the 12,333 women who had at least four ANC visits, regardless of the timing of their first visit, reported being late for ANC.

**CONCLUSION:** Early ANC4+ uptake increased in Zambia between 2007 and 2019. There were, however, disparities due to wealth, education, and parity. We found that 27% of women who were misclassified as having at least ANC4+ using conventional analysis were actually late for ANC. We provide some key considerations for ensuring that Zambia and other similar settings achieve universal antenatal care coverage by 2030.

## INTRODUCTION

Several antenatal care (ANC) models have been implemented by the World Health Organisation (WHO) since 2000 [1]. For example, the Focused Antenatal Care (FANC), an ANC model that recommends least four ANC (ANC4+) visits for women with uncomplicated pregnancies, has been implemented since 2000 [2]. Despite the reduction in maternal and child mortality due to improved health system in the past decade, most women still do not complete the required ANC visits since they report late for their first ANC visit [3]. Although most women under the FANC model did not meet the required minimal number of ANC visits, the WHO 2016 ANC guidelines doubled the number of ANC visits to be completed by ANC women [4] [5]. The WHO 2016 ANC guidelines are premised on high income countries where the increased number of contact has been associated with low perinatal and maternal deaths [5] [6].

There is evidence on the general uptake and timing of ANC services in Zambia [7] [8] [9] however such evidence have not assessed the composition of both early initiation and attendance of at least four ANC visits. Furthermore, in Zambia nearly half of the women had at least four ANC visits irrespective of the timing of the first ANC visit and that most of women report late for ANC [7] [8] [9]. This study, therefore, generates evidence that will allow policy makers and decision makers in Zambia and other similar settings to understand the determinants of both uptake and timing of ANC visits.

Furthermore, the evidence is important in informing the design and implementation of interventions on promoting both early initiation and attainment of the minimum of four ANC visits, and consequently informing the possibility of achieving the 8 contacts as stipulated in the WHO 2016 ANC guidelines.

## METHODS

### Study design, participants, data source, and sample size

We combined data from the 2007, 2013-4 and 2019 ZDHS. A secondary analysis of the women’s questionnaire data from three Zambia Demographic and Health Surveys (ZDHS) administered between 2007 and 2019 [10] [11]. These ZDHSs collected data from female participants on maternal and child health and healthcare use, contraception and women’s socio-economic status. All 20,661 women aged between 15-49 years who had given at least one birth during the five-year period preceding each survey were included in the analysis.

### Study setting

Zambia is located in southern Africa. Currently, there were approximately 19 million persons in Zambia in 2021. Zambia shares its border with Angola, Botswana, the Democratic Republic of Congo, Malawi, Mozambique, Namibia, Tanzania, and Zimbabwe. The country is divided into 116 districts and 10 provinces. The ANC services are provided free of user fees in all public health clinics.

### Sampling Procedure

The sampling frame for 2007 ZDHS was the Census of Population and Housing of the Republic of Zambia (CPH) conducted in 2000 [10] while the sampling frame for the 2014 and 2018 ZDHS was the 2010 Census of Population and Housing of the Republic of Zambia (CPH) [12]. The sampling frame is a complete list of all census standard enumeration areas (SEA). The SEA contains information about the location, type of residence (urban or rural), and the estimated number of residential households. The ZDHS samples were stratified and selected in two stages. A total of 320 SEAs were selected with probability proportional to size in the first stage. In the second stage, 300 households were selected. Prior to selection of the households, listing of the households in the selected clusters was done.

### Variables and Measurements

#### Outcome Variable

The outcome variable of interest was whether or not women had at least four ANC visits; reviewed by a doctor/clinical officer, or assistant clinical officer, or nurse/midwife; with the first visit occurring in or prior to the four months of pregnancy [13].

#### Independent Variable

The following were the explanatory variables used in this analysis and have been classified as follows: external environment (area of residence (rural/urban), year of interview, and province of residence during the interview), socio-demographics (woman’s age, wealth index of the household from which the woman comes from, woman’s highest education level, woman’s marital status, and number of children ever born by the woman), woman’s exposure to knowledge (frequency of listening to radio or watching television)) and woman’s ANC enablers (permission to visit health facility, availability of money to pay for health services, distance to health facilities, presence of companion when going for healthcare at a facility, and desire for current pregnancy) [13]. We further examined the health services accessed during ANC (blood test, urine test, blood pressure check-up, Iron tablets for 90+ days, HIV testing and counselling, and sulfadoxine-pyrimethamine (SP)/Fansidar for malaria prophylaxis) [13].

#### Data Management and statistical Analysis

Statistical data management and analysis were done in Stata version 17 (Stata Corp., Texas, USA). To detect bias, we examined the data for completeness and all the merged data had no missing data hence all the data were included in this analysis. We estimated frequencies, percentages, odds ratios (OR) and their associated 95% confidence intervals (95%CI) as applicable. We weighted the analysis since it came from a complex survey design [14]. We calculated the equal weights for each sample cluster and divided the average weight for each cluster by three as illustrated by Friedman and Jang in 2002 [15]. We then conducted weighted univariate, bivariate and multivariable logistic regression analysis of the effects of each of the explanatory variable on the binary endpoint of early initiation of ANC4 [13]. Multiple weighted logistic regression models were used with a forward step-wise selection method. We used the likelihood ratio test (LRT) to determine whether a factor was included in the model or not. We set a statistical significance of P< 0.05 as the threshold in this analysis. We included age of the woman *apriori* in the multiple logistic regression model.

#### Ethical Considerations

We obtained permission to conduct secondary analysis of the DHS 2007, 2013-14 and 2018 from Measure DHS. The ZDHS data were obtained data from Measure DHS. Individual informed consent was not required since we conducted secondary data analysis.

## RESULTS

### Characteristics of ANC women in Zambia

The characteristics of 20661 ANC women evaluated between 2007 and 2019 are shown in Table 1. A total of 4099 (20% of 20661) were interviewed in 2007 while 7305 (36% of 20661) were interviewed in the 2018/19. Table 1 shows that the majority of the women being aged 20-24 years while the minority were aged between 45 to 49 years. The median age of the ANC women was 28 years (interquartile range (IQR): 23-34). The majority of the ANC women had between 2 and 3 children previously (33%) while the minority of the ANC women had one child (22%) as shown in Table 1. The median parity was 3 (IQR: 2-5). Most of women had primary education while the minority had tertiary education (see Table 1). Forty-five percent of 20661 ANC women were from households of poor socio-economic position.

**Table 1:** Characteristics of women Interviewed during the Zambia Demographic and Health Surveys Conducted between 2007 and 2019.

| Characteristics (n =20661) | Total |  | 2007 |  | 2013-14 |  | 2018-19 |  |
| --- | --- | --- | --- | --- | --- | --- | --- | --- |
|  | n | % | N | % | n | % | n | % |
| **Total** | 20661 | 100.0 | 4099 | 100.0 | 9257 | 100.0 | 7305 | 100.0 |
| **Age group** |  |  |  |  |  |  |  |  |
| 15-19 | 1939 | 9.3 | 361 | 8.7 | 848 | 9.0 | 730 | 10.0 |
| 20-24 | 5067 | 24.4 | 1042 | 25.3 | 2139 | 23.0 | 1886 | 25.6 |
| 25-29 | 4996 | 24.3 | 1103 | 26.8 | 2262 | 24.5 | 1631 | 22.6 |
| 30-34 | 4005 | 19.6 | 782 | 19.2 | 1865 | 20.6 | 1358 | 18.5 |
| 35-39 | 2755 | 13.5 | 476 | 11.8 | 1280 | 13.8 | 999 | 14.0 |
| 40-44 | 1468 | 7.0 | 246 | 6.1 | 679 | 7.1 | 543 | 7.2 |
| 45-49 | 431 | 2.0 | 89 | 2.2 | 184 | 2.0 | 158 | 2.0 |
| **Number of children ever born** |  |  |  |  |  |  |  |  |
| 1 | 4481 | 21.7 | 797 | 19.2 | 1890 | 20.4 | 1794 | 24.8 |
| 2-3 | 6788 | 33.3 | 1352 | 33.2 | 2996 | 32.9 | 2440 | 33.8 |
| 4-5 | 4699 | 22.8 | 988 | 24.1 | 2144 | 23.3 | 1567 | 21.4 |
| 6+ | 4693 | 22.2 | 962 | 23.5 | 2227 | 23.4 | 1504 | 20.0 |
| **Education level** |  |  |  |  |  |  |  |  |
| None | 2199 | 10.4 | 520 | 12.7 | 954 | 10.1 | 725 | 9.6 |
| Primary | 11055 | 53.5 | 2459 | 60.1 | 4924 | 53.6 | 3672 | 49.5 |
| Secondary | 6587 | 32.1 | 1004 | 24.3 | 3004 | 32.1 | 2579 | 36.5 |
| Tertiary | 820 | 4.0 | 116 | 2.8 | 375 | 4.2 | 329 | 4.4 |
| **Wealth index quintile** |  |  |  |  |  |  |  |  |
| Poorest | 4750 | 22.5 | 834 | 20.4 | 2021 | 22.1 | 1895 | 24.1 |
| Poorer | 4540 | 21.2 | 829 | 20.1 | 2033 | 21.1 | 1678 | 22.0 |
| Middle | 4523 | 20.9 | 909 | 21.0 | 2134 | 21.6 | 1480 | 19.9 |
| Richer | 3839 | 19.5 | 935 | 22.9 | 1718 | 19.0 | 1186 | 18.2 |
| Richest | 3009 | 15.9 | 592 | 15.6 | 1351 | 16.2 | 1066 | 15.7 |
| **Residence** |  |  |  |  |  |  |  |  |
| Urban | 7608 | 37.6 | 1460 | 35.8 | 3747 | 39.7 | 2401 | 36.1 |
| Rural | 13053 | 62.4 | 2639 | 64.2 | 5510 | 60.3 | 4904 | 63.9 |
| **Sources of antenatal care knowledge** |  |  |  |  |  |  |  |  |
| **Frequency of listening to radio** |  |  |  |  |  |  |  |  |
| Less than once a week | 11670 | 56.4 | 1868 | 46.3 | 4791 | 51.6 | 5011 | 68.2 |
| At least once a week | 8986 | 43.6 | 2231 | 53.7 | 4461 | 48.4 | 2294 | 31.8 |
| **Frequency of watching television** |  |  |  |  |  |  |  |  |
| Less than once a week | 15163 | 71.5 | 3179 | 76.8 | 6549 | 68.7 | 5435 | 72.1 |
| At least once a week | 5498 | 28.5 | 920 | 23.2 | 2708 | 31.3 | 1870 | 27.9 |
| **Barriers to access antenatal care** |  |  |  |  |  |  |  |  |
| **Permission to visit health services** |  |  |  |  |  |  |  |  |
| Big problem | 803 | 3.8 | 181 | 4.5 | 283 | 3.0 | 339 | 4.5 |
| Not big problem | 19858 | 96.2 | 3918 | 95.5 | 8974 | 97.0 | 6966 | 95.5 |
| **Money to pay for health services** |  |  |  |  |  |  |  |  |
| Big problem | 5585 | 26.7 | 1360 | 34.4 | 2494 | 26.3 | 1731 | 22.9 |
| Not big problem | 15076 | 73.3 | 2739 | 65.6 | 6763 | 73.7 | 5574 | 77.1 |
| **Distance to health facilities** |  |  |  |  |  |  |  |  |
| Big problem | 8180 | 38.6 | 1775 | 43.5 | 3882 | 41.3 | 2523 | 32.5 |
| Not big problem | 12481 | 61.4 | 2324 | 56.5 | 5375 | 58.7 | 4782 | 67.5 |
| **Presence of companion** |  |  |  |  |  |  |  |  |
| Big problem | 3897 | 18.4 | 1099 | 26.7 | 1669 | 18.1 | 1129 | 14.1 |
| Not big problem | 16758 | 81.6 | 3000 | 73.3 | 7582 | 81.9 | 6176 | 85.9 |
| **No female provider** |  |  |  |  |  |  |  |  |
| Big problem | 2629 | 12.5 | 788 | 18.4 | 985 | 10.8 | 856 | 11.4 |
| Not big problem | 18024 | 87.5 | 3311 | 81.6 | 8264 | 89.2 | 6449 | 88.6 |
| **Marital status** |  |  |  |  |  |  |  |  |
| Never married | 2431 | 11.4 | 398 | 9.3 | 1019 | 10.2 | 1014 | 14.0 |
| Married | 15952 | 77.8 | 3280 | 80.2 | 7223 | 79.1 | 5449 | 74.7 |
| Widowed | 419 | 2.0 | 110 | 2.8 | 196 | 2.0 | 113 | 1.5 |
| Divorced | 1859 | 8.8 | 311 | 7.7 | 819 | 8.6 | 729 | 9.7 |

Sixty-two percent of the women were from rural areas with more women were being from rural areas between 2007 and 2019 (see Table 1). Although 8986 of 20661 listened to radio between 2007 and 2019, we observed a decreasing trend from 54% in 2007 to 32% in 2018/19. Overall, the minority of women (5498 of 20661) watched a television (TV) for more than once a week but the numbers watching TV increased from 23% in 2007 to 28% in 2018/19. Over the 2007 to 2019 surveys, women cited different barriers to accessing ANC and these barriers had different trends (see Table 1). The major barriers were long distance to health facilities (39% of 20661) and lack of money to use in accessing health services (27% of 20661). Across the survey populations, the majority of the women were married while the lowest proportion were widowed (see Table 1).

### Factors associated with early antenatal care of at least four visits in Zambia

#### Distribution of women by number of ANC visits

The distribution of women by number of ANC visits is shown in Table 2a and 2b. Of the 20661, 11633 (56%) of women had attended early of the ANC4+. There was an increasing trend in the uptake of early initiation of ANC4+ from 55% in 2007 to 63% in 2018/19 (P<0.001). The distribution of women with early initiation of ANC4+ decreased with increasing parity (see Table 2a). Early ANC+ initiation was higher amongst women with tertiary education (74%) compared to those with no formal education (52%). Also, early initiation of ANC4+ was higher amongst women from rural areas than their counterparts from urban areas (see Table 2a).

**Table 2a:** Distribution of women with at least four ANC visits in Zambia between 2007 and 2019.

| Characteristics | Total |  |  |  | 2007 |  |  |  | 2013-14 |  |  |  | 2018-19 |  |  |  |
| --- | --- | --- | --- | --- | --- | --- | --- | --- | --- | --- | --- | --- | --- | --- | --- | --- |
|  | <4 ANC |  | ANC4+ |  | <4 ANC |  | ANC4+ |  | <4 ANC |  | ANC4+ |  | <4 ANC |  | ANC4+ |  |
|  | n | % | n | % | n | % | N | % | n | % | n | % | n | % | n | % |
| **Total** | 9028 | 43.6 | 11633 | 56.4 | 1848 | 45.1 | 2251 | 54.9 | 4459 | 47.8 | 4798 | 52.2 | 2721 | 37.5 | 4584 | 62.5 |
| **Age group** |  |  |  |  |  |  |  |  |  |  |  |  |  |  |  |  |
| 15-19 | 901 | 46.6 | 1038 | 53.4 | 180 | 48.7 | 181 | 51.3 | 433 | 51.5 | 415 | 48.5 | 288 | 40.1 | 442 | 59.9 |
| 20-24 | 2285 | 45.1 | 2782 | 54.9 | 487 | 47.1 | 555 | 52.9 | 1090 | 51.2 | 1049 | 48.8 | 708 | 37.2 | 1178 | 62.8 |
| 25-29 | 2202 | 43.6 | 2794 | 56.4 | 479 | 42.7 | 624 | 57.3 | 1102 | 47.5 | 1160 | 52.5 | 621 | 38.8 | 1010 | 61.2 |
| 30-34 | 1694 | 42.2 | 2311 | 57.8 | 330 | 42.4 | 452 | 57.6 | 881 | 46.8 | 984 | 53.2 | 483 | 35.6 | 875 | 64.4 |
| 35-39 | 1133 | 41.1 | 1622 | 58.9 | 208 | 44.5 | 268 | 55.5 | 566 | 43.3 | 714 | 56.7 | 359 | 36.8 | 640 | 63.2 |
| 40-44 | 623 | 43.0 | 845 | 57.0 | 116 | 48.1 | 130 | 51.9 | 301 | 45.4 | 378 | 54.6 | 206 | 37.7 | 337 | 62.3 |
| 45-49 | 190 | 45.2 | 241 | 54.8 | 48 | 57.5 | 41 | 42.5 | 86 | 46.6 | 98 | 53.4 | 56 | 35.7 | 102 | 64.3 |
| **Number of children ever born** |  |  |  |  |  |  |  |  |  |  |  |  |  |  |  |  |
| 1 | 1890 | 42.0 | 2591 | 58.0 | 344 | 42.1 | 453 | 57.9 | 890 | 46.8 | 1000 | 53.2 | 656 | 37.0 | 1138 | 63.0 |
| 2-3 | 2995 | 43.8 | 3793 | 56.2 | 605 | 44.7 | 747 | 55.3 | 1485 | 48.9 | 1511 | 51.1 | 905 | 37.2 | 1535 | 62.8 |
| 4-5 | 2031 | 43.3 | 2668 | 56.7 | 435 | 43.7 | 553 | 56.3 | 1034 | 48.2 | 1110 | 51.8 | 562 | 36.4 | 1005 | 63.6 |
| 6+ | 21[12 | 45.2 | 2581 | 54.8 | 464 | 49.6 | 498 | 50.4 | 1050 | 46.7 | 1177 | 53.3 | 598 | 40.1 | 906 | 59.9 |
| **Education level** |  |  |  |  |  |  |  |  |  |  |  |  |  |  |  |  |
| None | 1060 | 48.2 | 1139 | 51.8 | 258 | 49.4 | 262 | 50.6 | 498 | 52.7 | 456 | 47.3 | 304 | 41.5 | 421 | 58.5 |
| Primary | 4873 | 44.3 | 6182 | 55.7 | 1119 | 46.0 | 1340 | 54.0 | 2386 | 48.4 | 2538 | 51.6 | 1368 | 37.5 | 2304 | 62.5 |
| Secondary | 2881 | 43.2 | 3706 | 56.8 | 442 | 43.1 | 562 | 56.9 | 1454 | 47.6 | 1550 | 52.4 | 985 | 38.5 | 1594 | 61.5 |
| Tertiary | 214 | 25.9 | 606 | 74.1 | 29 | 24.1 | 87 | 75.9 | 121 | 30.5 | 254 | 69.5 | 64 | 21.0 | 265 | 79.0 |
| **Wealth index quintile** |  |  |  |  |  |  |  |  |  |  |  |  |  |  |  |  |
| Poorest | 2093 | 44.5 | 2657 | 55.5 | 398 | 48.3 | 436 | 51.7 | 1010 | 50.6 | 1011 | 49.4 | 685 | 35.8 | [1210 | 64.2 |
| Poorer | 1907 | 42.3 | 2633 | 57.7 | 352 | 43.1 | 477 | 56.9 | 963 | 47.5 | 1070 | 52.5 | 592 | 35.8 | 1086 | 64.2 |
| Middle | 2028 | 44.2 | 2495 | 55.8 | 397 | 42.4 | 5[12 | 57.6 | 1058 | 49.1 | 1076 | 50.9 | 573 | 38.7 | 907 | 61.3 |
| Richer | 1844 | 47.7 | 1995 | 52.3 | 454 | 48.6 | 481 | 51.4 | 880 | 51.1 | 838 | 48.9 | 510 | 42.7 | 676 | 57.3 |
| Richest | 1156 | 38.2 | 1853 | 61.8 | 247 | 42.2 | 345 | 57.8 | 548 | 38.9 | 803 | 61.1 | 361 | 35.1 | 705 | 64.9 |
| **Residence** |  |  |  |  |  |  |  |  |  |  |  |  |  |  |  |  |
| Urban | 3535 | 45.4 | 4073 | 54.6 | 674 | 45.7 | 786 | 54.3 | 1874 | 48.7 | 1873 | 51.3 | 987 | 40.8 | 1414 | 59.2 |
| Rural | 5493 | 42.5 | 7560 | 57.5 | 1174 | 44.8 | 1465 | 55.2 | 2585 | 47.2 | 2925 | 52.8 | 1734 | 35.7 | 3170 | 64.3 |

**Table 2b:** Distribution of women with at least four ANC visits in Zambia between 2007 and 2019.

| Characteristics | Total |  |  |  | 2007 |  |  |  | 2013-14 |  |  |  | 2018-19 |  |  |  |
| --- | --- | --- | --- | --- | --- | --- | --- | --- | --- | --- | --- | --- | --- | --- | --- | --- |
|  | <4 ANC |  | ANC4+ |  | <4 ANC |  | ANC4+ |  | <4 ANC |  | ANC4+ |  | <4 ANC |  | ANC4+ |  |
|  | n | % | n | % | n | % | N | % | n | % | n | % | n | % | n | % |
| **Sources of antenatal care knowledge** |  |  |  |  |  |  |  |  |  |  |  |  |  |  |  |  |
| **Frequency of listening to radio** |  |  |  |  |  |  |  |  |  |  |  |  |  |  |  |  |
| Less than once a week | 5095 | 44.0 | 6575 | 56.0 | 862 | 46.2 | 1006 | 53.8 | 2337 | 49.1 | 2454 | 50.9 | 1896 | 38.3 | 3115 | 61.7 |
| At least once a week | 3928 | 43.1 | 5058 | 56.9 | 986 | 44.2 | 1245 | 55.8 | 2117 | 46.4 | 2344 | 53.6 | 825 | 35.9 | 1469 | 64.1 |
| **Frequency of watching television** |  |  |  |  |  |  |  |  |  |  |  |  |  |  |  |  |
| Less than once a week | 6688 | 44.2 | 8475 | 55.8 | 1448 | 45.7 | 1731 | 54.3 | 3206 | 49.1 | 3343 | 50.9 | 2034 | 37.4 | 3401 | 62.6 |
| At least once a week | 2340 | 42.2 | 3158 | 57.8 | 400 | 43.2 | 520 | 56.8 | 1253 | 45.0 | 1455 | 55.0 | 687 | 37.9 | 1183 | 62.1 |
| **Barriers to access antenatal care** |  |  |  |  |  |  |  |  |  |  |  |  |  |  |  |  |
| **Permission to visit health services** |  |  |  |  |  |  |  |  |  |  |  |  |  |  |  |  |
| Big problem | 393 | 49.4 | 410 | 50.6 | 96 | 52.5 | 85 | 47.5 | 148 | 55.1 | 135 | 44.9 | 149 | 43.1 | 190 | 56.9 |
| Not big problem | 8635 | 43.4 | 11223 | 56.6 | 1752 | 44.8 | 2166 | 55.2 | 4311 | 47.6 | 4663 | 52.4 | 2572 | 37.3 | 4394 | 62.7 |
| **Money to pay for health services** |  |  |  |  |  |  |  |  |  |  |  |  |  |  |  |  |
| Big problem | 2529 | 45.0 | 3056 | 55.0 | 658 | 48.2 | 702 | 51.8 | 1216 | 48.2 | 1278 | 51.8 | 655 | 37.9 | 1076 | 62.1 |
| Not big problem | 6499 | 43.1 | 8577 | 56.9 | 1190 | 43.5 | 1549 | 56.5 | 3243 | 47.7 | 3520 | 52.3 | 2066 | 37.4 | 3508 | 62.6 |
| **Distance to health facilities** |  |  |  |  |  |  |  |  |  |  |  |  |  |  |  |  |
| Big problem | 3678 | 45.3 | 4502 | 54.7 | 841 | 47.4 | 934 | 52.6 | 1861 | 48.5 | 2021 | 51.5 | 976 | 38.5 | 1547 | 61.5 |
| Not big problem | 5350 | 42.6 | 7131 | 57.4 | 1007 | 43.4 | 1317 | 56.6 | 2598 | 47.3 | 2777 | 52.7 | 1745 | 37.0 | 3037 | 63.0 |
| **Presence of companion** |  |  |  |  |  |  |  |  |  |  |  |  |  |  |  |  |
| Big problem | 1745 | 44.5 | 2152 | 55.5 | 491 | 44.2 | 608 | 55.8 | 824 | 48.9 | 845 | 51.1 | 430 | 37.6 | 699 | 62.4 |
| Not big problem | 7277 | 43.4 | 9481 | 56.6 | 1357 | 45.5 | 1643 | 54.5 | 3629 | 47.5 | 3953 | 52.5 | 2291 | 37.5 | 3885 | 62.5 |
| **No female provider** |  |  |  |  |  |  |  |  |  |  |  |  |  |  |  |  |
| Big problem | 1163 | 44.1 | 1466 | 55.9 | 355 | 44.9 | 433 | 55.1 | 459 | 46.4 | 526 | 53.6 | 349 | 40.8 | 507 | 59.2 |
| Not big problem | 7857 | 43.5 | 10167 | 56.5 | 1493 | 45.2 | 1818 | 54.8 | 3992 | 47.9 | 4272 | 52.1 | 2372 | 37.1 | 4077 | 62.9 |
| **Marital status** |  |  |  |  |  |  |  |  |  |  |  |  |  |  |  |  |
| Never married | 1111 | 54.2 | 1320 | 45.8 | 190 | 47.7 | 208 | 52.3 | 525 | 51.4 | 494 | 48.6 | 396 | 39.9 | 618 | 60.1 |
| Married | 6911 | 43.3 | 9041 | 56.7 | 1493 | 45.6 | 1787 | 54.4 | 3433 | 47.2 | 3790 | 52.8 | 1985 | 36.6 | 3464 | 63.4 |
| Widowed | 179 | 43.9 | 240 | 56.1 | 44 | 42.0 | 66 | 58.0 | 89 | 47.4 | 107 | 52.6 | 46 | 39.9 | 67 | 60.1 |
| Divorced | 827 | 43.9 | 1032 | 56.1 | 121 | 38.4 | 190 | 61.6 | 412 | 49.3 | 407 | 50.7 | 294 | 40.4 | 435 | 59.6 |

#### Adjusted odds ratios of women having early antenatal care of at least four visits

Table 3 shows both the crude and adjusted odds ratios for early initiation of ANC4+ amongst women in Zambia. The uptake of early initiation of ANC4+ significantly increased between 2007 and 2019 (P<0.001). However, the odds of early initiation of ANC4+ decreased with increasing parity (see Table 3) while there was an increasing uptake of early initiation of ANC4+ with the level of education (Table 3). Women from richer households were less likely to have early initiation of ANC4+ compared to their counterparts from poor households (OR= 0.81, 95%CI: 0.66-0.99, P=0.03).

**Table 3:**
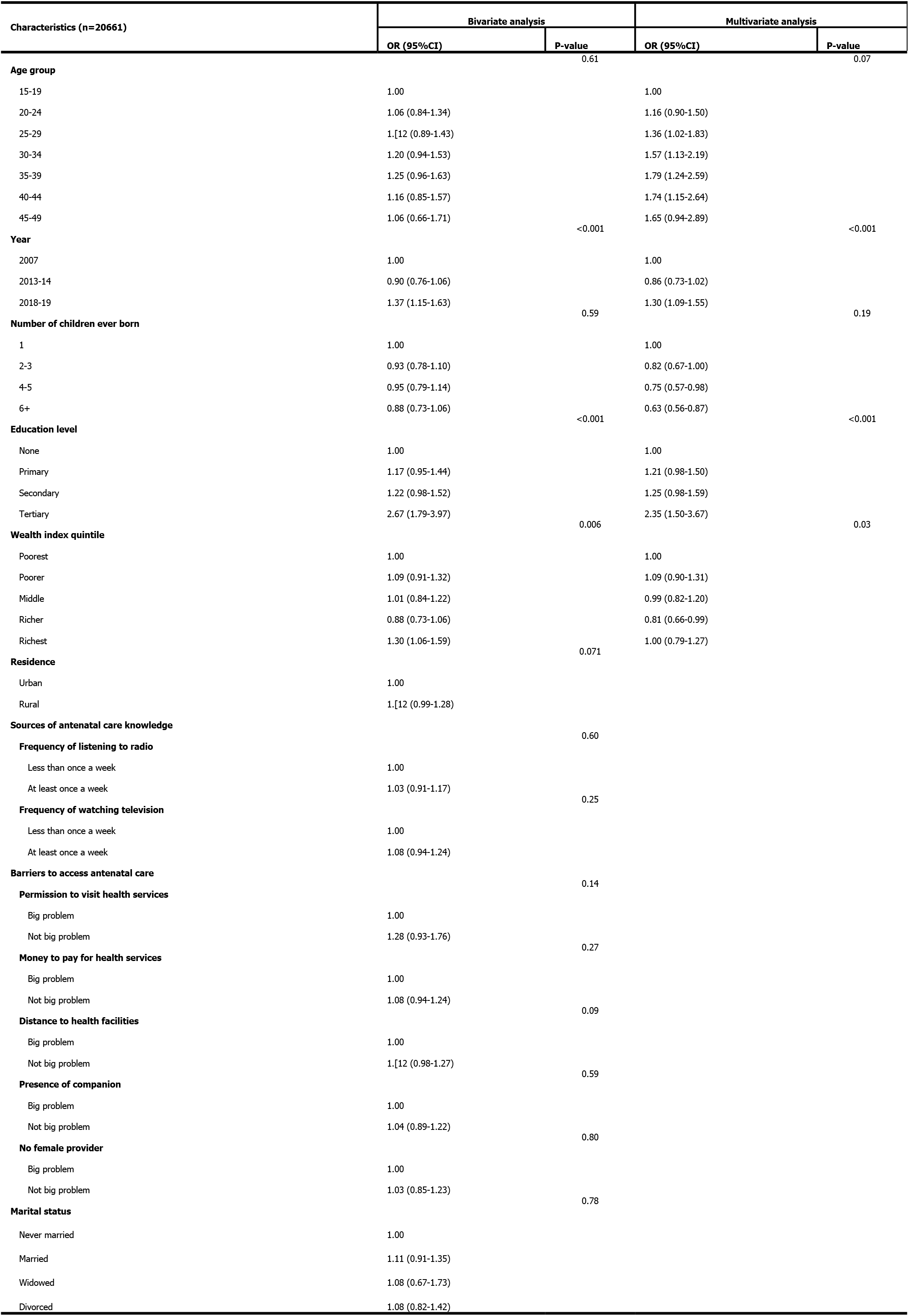
Bivariate and multivariate odds ratios for factors associated with four or more antenatal care visits in Zambia between 2007 and 2019.

#### Services received by women seeking antenatal care

Figure 1 shows the ANC services received by women that attended ANC in Zambia. Between 2007 and 2019, the coverage of full blood count increased from 61% to 96% (P <0.001). Similarly, urine test was done 23% of the ANC women in 2007 and 64% of the ANC women in 2018/19 (P<0.001). The distribution of ANC women that were given at least two doses of Fansidar/SP for malaria prophylaxis increased from 86% in 2007 to 95% in 2018/19 (P<0.001). HIV testing uptake at ANC increased from 46% in 2007 to 70% in 2013/14 and then dropped to 67% in 2018/19. Between 2007 and 2019, blood pressure checkup increased from 81% to 94% of the ANC women. By 2019, nearly all ANC women in Zambia received iron tablets (see Figure 1).

**Figure 1.**
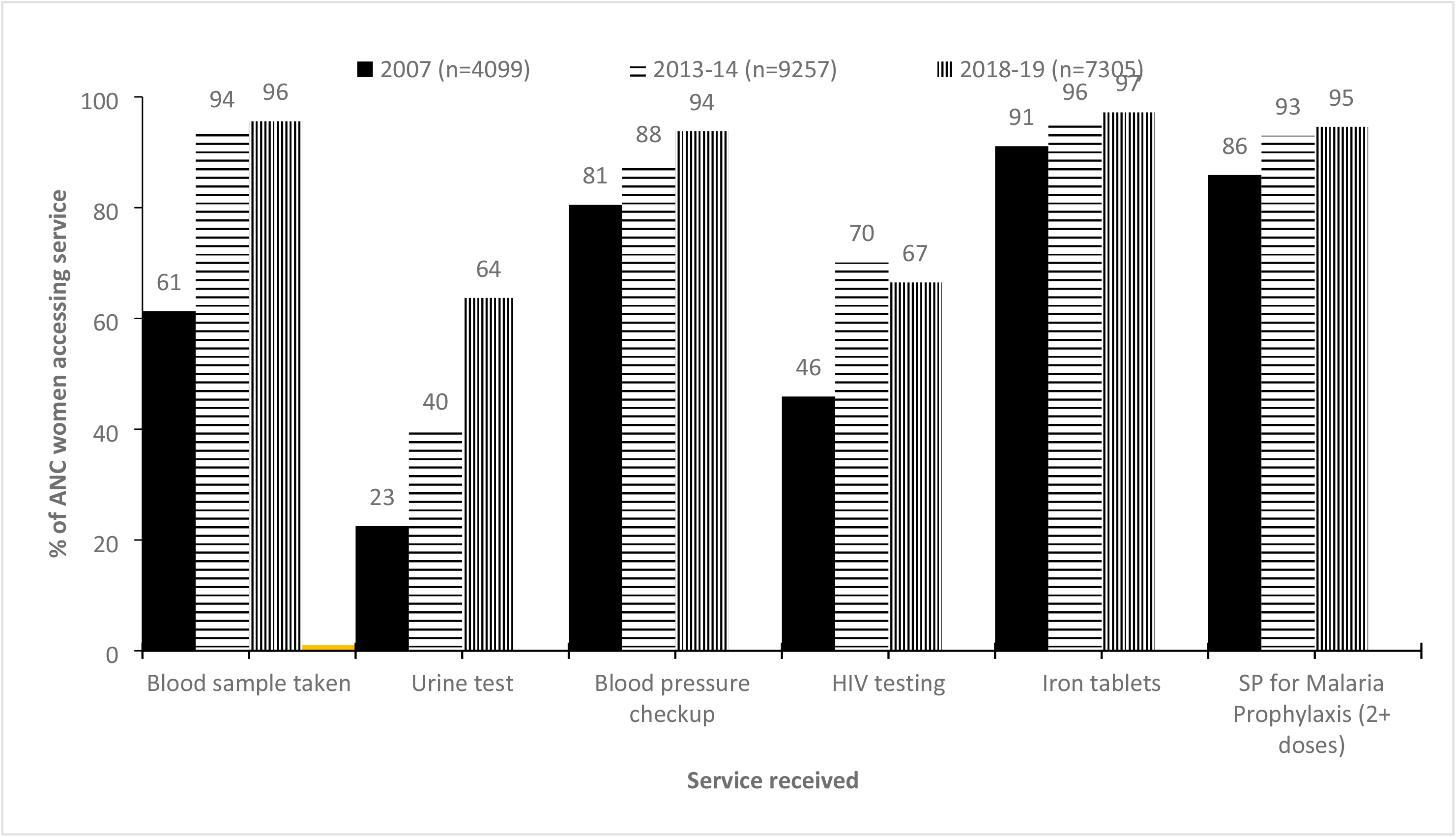
Services received by women that attended antenatal care in Zambia between 2007 and 2019.

#### Timing of antenatal care visits

The median time of presenting for the first ANC was 4 months (IQR: 3-5). Over the time period, there was strong evidence of association between timing of first ANC visit by survey year (P<0.001). Table 4 shows the characteristics of women with at least four ANC visits regardless of the timing of the first ANC visit. Of the 12333 ANC women with at least 4 ANC visits, 9004 (73%) started ANC after the fourth month of pregnancy. The distribution of ANC women that attended their first ANC visit after four months dropped from 40% in 2007 to 19% in 2018/19. Most women with higher parity had late attendance of ANC (see Table 4). In addition, fewer ANC women with tertiary education (21%) than women with no formal education (26%) had late attendance of ANC.

**Table 4:**
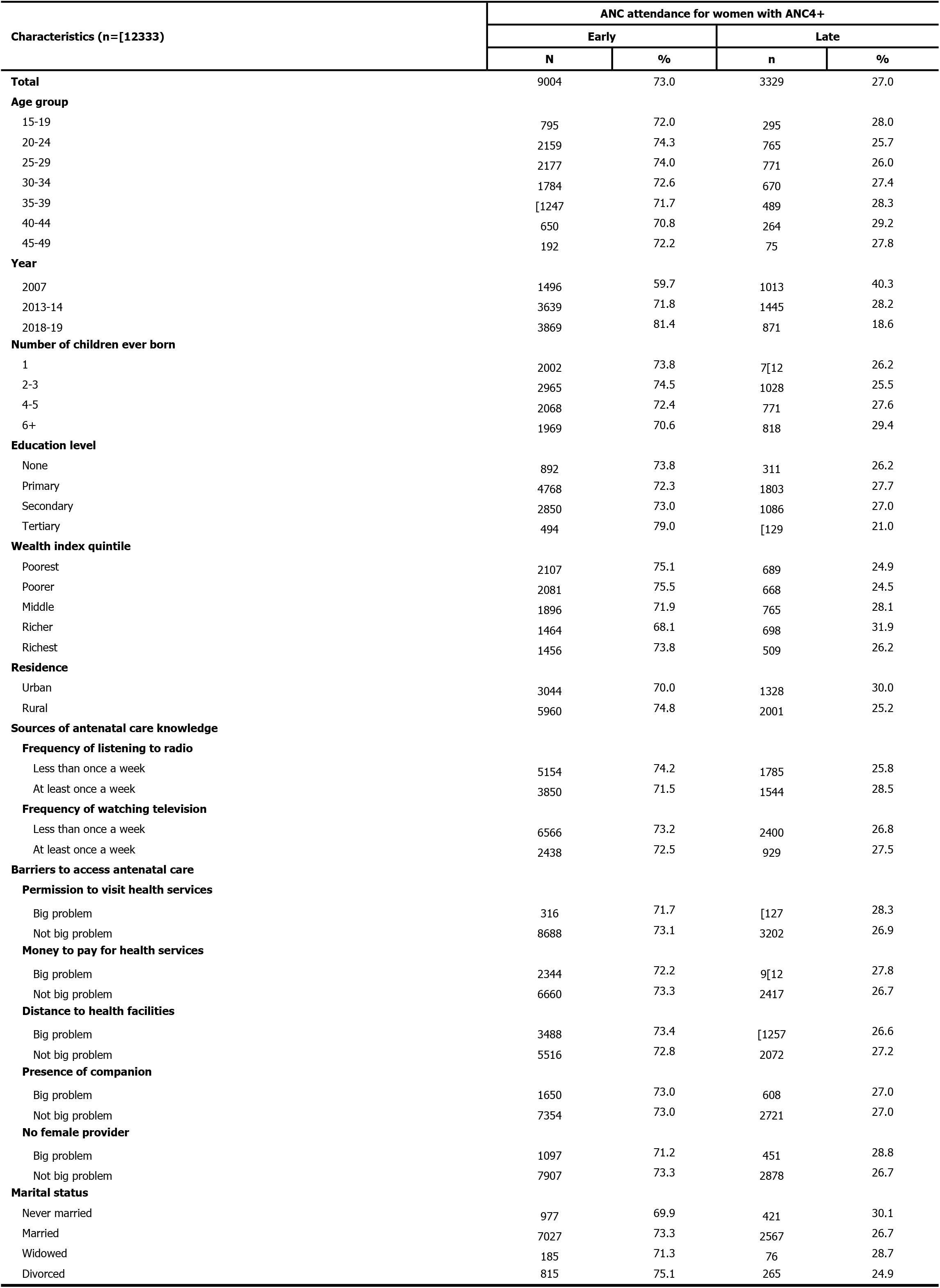
Distribution of women with at least four antenatal care visits by socio-demographic characteristics of the women in Zambia between 2007 and 2019.

## DISCUSSION

This is the first analysis of both early initiations and uptake of four or more ANC visits through the using combined outcome in the context of Zambia between 2007 and 2019. The study’s key findings were: (a) nearly two-thirds of the women in Zambia had early ANC4+ visits, (b) there was increasing trend in early ANC4+ visits between 2007 and 2019, (c) early of ANC4+ decreased with increasing parity, (d) women from richer households were less likely to have early ANC4+, (d) early of ANC4+ increased with increasing level of woman’s education, and (e) uptake of services provided to ANC women increased although HIV testing was sub-optimal.

Our study The uptake of early ANC4+ in Zambia was 56%. The observed uptake of early ANC4+ for Zambia was almost twice the early ANC4+ visits reported in Malawi over similar time period [13]. The difference in the uptake of early ANC4+ is attributable to the existing policy frameworks in operationalizing the ANC guidelines to achieve the targets for the two countries [7] [16].

We observed an increasing trend in early ANC4+ uptake over time between 2007 and 2019 for Zambia. The observed trend in early ANC4+ is similar to what was observed in Malawi [13]. Similarly, a multi-country analysis of the countries in the East Africa region found increasing uptake of ANC4+ visits over time [17]. The increasing uptake of early ANC4+ is mainly due to increased commitment of these countries in implementing the global and local interventions aimed at providing quality ANC4+ services to women in these settings [18]. In Zambia, the emphasis was placed on empowering community volunteers also known as Safe Motherhood Action Groups (SMAGs) with health promotion messages to encourage women to report early for ANC in order to achieve ANC4+ visits and access health facility delivery [19] [20].

In this study we found a lower uptake of early ANC4+ with increasing parity. Other similar studies have also observed a similar trend in uptake of early ANC4+ with increasing parity [17] [13]. A possible explanation for the observed pattern could be that women who have more children feel more experienced with pregnancy due to previous experiences with non-complicated pregnancies and end up reporting late for ANC. In addition, women with a higher parity may also be ashamed with their current pregnancies and hence find themselves not willing to report early for ANC and this is mainly the case with unwanted pregnancies or multiparty.

We also observed that women from richer households were less likely to have early ANC4+ than the women from poorest households. The findings from this study are different from what has been observed in sub-Saharan Africa (SSA) [21] or in Malawi [13]. The differences may be attributable to different socio-cultural environments in which the ANC guidelines are implemented, where deliberate efforts are placed on poor communities to the neglect of those living in richer communities. Such disparities can be addressed by embracing ideologies for universal coverage for all health care services and especially reproductive health interventions [22].

In this study, there was an increasing uptake of early ANC4+ with increasing education level of the ANC women. This is similar to what was observed in Malawi on a study that looked at the uptake of early ANC4+ [13]. Furthermore, a multi-country study involving 36 countries in SSA also found huge inequalities in uptake of ANC by woman’s highest level of education in which those with lower education had lower uptake of ANC [21]. Therefore, Zambia should potentially invest in education so that disparities that occur as a result of lower education get reduced or eliminated completely. This would also contribute towards achieving universal health coverage of the ANC and other health outcomes that are sensitive to education.

There has been an increasing trend in the services offered within the ANC setting in Zambia except for HIV testing. However, other settings like Malawi [13] and Uganda [23] have higher rates of HIV testing in ANC than what has been observed in Zambia. Needless to mention that ANC is one of the critical service delivery points for identifying HIV positive women in the prevention and elimination mother to child transmission [23] [24] [25] [26] [27]. The observed HIV testing uptake amongst ANC women in Zambia was quite below the UNAIDS 95% target for HIV testing [28]. There is need to strategies for integrated HIV testing in ANC services in order to move towards universal HIV testing in antenatal care settings.

The main strength of the current approach to analyzing the ANC4+ visits is that it minimizes the risk of over-reporting the women with ANC4+ visits. In this study, 27% of the women that would have been misclassified as having at least 4 ANC visits actually reported late for ANC and their frequent visits were due to pregnancy related complications. Some of the limitations in this study are (a) that the analysis does not include data on quality of the ANC services received by the women in Zambia, (b) although hypertension, diabetes and previous HIV status prior to the current pregnancy might affect ANC uptake, the ZDHS did not capture these hence they were not included in this analysis, and (c) although ZDHS collected information on HIV testing, there is no information on the timing of the captured HIV testing to the pregnancy.

## CONCLUSION

In conclusion, early uptake of ANC4+ increased between 2007 and 2019 in Zambia. However, there were inequalities due to wealth, education and parity. Our current analytical approach has demonstrated that 27% of the women that would have been misclassified as having at least ANC4+ using conventional analysis were actually late for ANC. The countries in Sub-Saharan Africa should put in place careful measures in implementing the 2016 ANC guidelines since most of them are not achieving at least four timely ANC visits has been a toll order. The findings provide some key considerations in ensuring that Zambia and other similar settings are able to achieve universal coverage of antenatal care by 2030.

## Data Availability

Data available within the attachments of the manuscript

https://www.dhsprogram.com/data/available-datasets.cfm

## AUTHORS’ CONTRIBUTIONS

WFN led the manuscript writing, conducted data management and analysis; CZ, ANH, ASM, DN, TH & AM advised on the data analysis and policy insights on the paper. All authors read and approved the final manuscript.

### WHAT IS ALREADY KNOWN ON THIS TOPIC

Most studies have analysed timing of ANC visits and uptake of ANC4+ visits separately.

### WHAT THIS STUDY ADDS

Generates evidence that will allow policy makers and decision makers in Zambia and other similar settings to understand the determinants of both uptake and timing of ANC visits as a combined outcome. This will be crucial for ANC programme planning and implementation.

## ACKNOWLEDGEMENT

The authors would like to thank the Measure DHS for allowing us to use the ZDHS data.

---

|  | Item No | Recommendation | Page No |
| --- | --- | --- | --- |
| 1Title and abstract | 1 | (a) Indicate the study’s design with a commonly used term in the title or the abstract | 1 |
|  |  | (b) Provide in the abstract an informative and balanced summary of what was done and what was found | 1 |
| Introduction |  |  |  |
| Background/rationale | 2 | Explain the scientific background and rationale for the investigation being reported | 2 |
| Objectives | 3 | State specific objectives, including any prespecified hypotheses | 3 |
| Methods |  |  |  |
| Study design | 4 | Present key elements of study design early in the paper | 3 |
| Setting | 5 | Describe the setting, locations, and relevant dates, including periods of recruitment, exposure, follow-up, and data collection | 3 |
| Participants | 6 | (a) Give the eligibility criteria, and the sources and methods of selection of participants | 3 |
| Variables | 7 | Clearly define all outcomes, exposures, predictors, potential confounders, and effect modifiers. Give diagnostic criteria, if applicable | 4 |
| Data sources/ measurement | 8* | For each variable of interest, give sources of data and details of methods of assessment (measurement). Describe comparability of assessment methods if there is more than one group | 4 |
| Bias | 9 | Describe any efforts to address potential sources of bias |  |
| Study size | 10 | Explain how the study size was arrived at | 3,5 |
| Quantitative variables | 11 | Explain how quantitative variables were handled in the analyses. If applicable, describe which groupings were chosen and why | 4 |
| Statistical methods | 12 | (a) Describe all statistical methods, including those used to control for confounding | 4 |
|  |  | (b) Describe any methods used to examine subgroups and interactions | 4 |
|  |  | (c) Explain how missing data were addressed |  |
|  |  | (d) If applicable, describe analytical methods taking account of sampling strategy | 4 |
|  |  | (e) Describe any sensitivity analyses |  |
| Results |  |  |  |
| Participants | 13* | (a) Report numbers of individuals at each stage of study—eg numbers potentially eligible, examined for eligibility, confirmed eligible, included in the study, completing follow-up, and analysed |  |
|  |  | (b) Give reasons for non-participation at each stage |  |
|  |  | (c) Consider use of a flow diagram |  |
| Descriptive data | 14* | (a) Give characteristics of study participants (eg demographic, clinical, social) and information on exposures and potential confounders | 5 |
|  |  | (b) Indicate number of participants with missing data for each variable of interest | 5 |
| Outcome data | 15* | Report numbers of outcome events or summary measures | 5, 6, 6 |
| Main results | 16 | (a) Give unadjusted estimates and, if applicable, confounder-adjusted estimates and their precision (eg, 95% confidence interval). Make clear which confounders were adjusted for and why they were included | 6 |
|  |  | (b) Report category boundaries when continuous variables were categorized |  |
|  |  | (c) If relevant, consider translating estimates of relative risk into absolute risk for a meaningful time period |  |
| Other analyses | 17 | Report other analyses done—eg analyses of subgroups and interactions, and sensitivity analyses |  |
| **Discussion** |  |  |  |
| Key results | 18 | Summarise key results with reference to study objectives | 7 |
| Limitations | 19 | Discuss limitations of the study, taking into account sources of potential bias or imprecision. Discuss both direction and magnitude of any potential bias |  |
| Interpretation | 20 | Give a cautious overall interpretation of results considering objectives, limitations, multiplicity of analyses, results from similar studies, and other relevant evidence |  |
| Generalisability | 21 | Discuss the generalisability (external validity) of the study results | 8 |
| **Other information** |  |  |  |
| Funding | 22 | Give the source of funding and the role of the funders for the present study and, if applicable, for the original study on which the present article is based | 10 |

\*Give information separately for exposed and unexposed groups.
**Note:** An Explanation and Elaboration article discusses each checklist item and gives methodological background and published examples of transparent reporting. The STROBE checklist is best used in conjunction with this article (freely available on the Web sites of PLoS Medicine at http://www.plosmedicine.org/, Annals of Internal Medicine at http://www.annals.org/, and Epidemiology at http://www.epidem.com/). Information on the STROBE Initiative is available at www.strobe-statement.org.

## REFERENCES

1. Villar J, Ba’aqeel H, Piaggio G, Lumbiganon P, Belizán JM, Farnot U, et al. WHO antenatal care randomised trial for the evaluation of a new model of routine antenatal care. The Lancet. 2001 May 19;357(9268):1551–64.

2. Mchenga M, Burger R, von Fintel D. Examining the impact of WHO’s Focused Antenatal Care policy on early access, underutilisation and quality of antenatal care services in Malawi: a retrospective study. BMC Health Serv Res. 2019 May 8;19(1):295.

3. Andegiorgish AK, Elhoumed M, Qi Q, Zhu Z, Zeng L. Determinants of antenatal care use in nine sub-Saharan African countries: a statistical analysis of cross-sectional data from Demographic and Health Surveys. BMJ Open. 2022 Feb 1;12(2):e051675.

4. WHO recommendations on antenatal care for a positive pregnancy experience [Internet]. Apps.who.int. 2016 [cited 12 August 2020]. Available from: https://apps.who.int/iris/bitstream/handle/10665/250796/9789241549912-eng.pdf?sequence=1.

5. World Health Organization (WHO). WHO Recommendations on Antenatal Care for a Positive Pregnancy Experience: Summary. Geneva, Switzerland: WHO; 2018. Licence: CC BY-NC-SA 3.0 IGO.

6. Vogel JP, Habib NA, Souza JP, Gülmezoglu AM, Dowswell T, Carroli G, et al. Antenatal care packages with reduced visits and perinatal mortality: a secondary analysis of the WHO Antenatal Care Trial. Reprod Health. 2013;10(1):1–7.

7. Chama-Chiliba CM, Koch SF. Utilization of focused antenatal care in Zambia: examining individual- and community-level factors using a multilevel analysis. Health Policy Plan. 2015 Feb 1;30(1):78–87.

8. Kyei NNA, Campbell OMR, Gabrysch S. The Influence of Distance and Level of Service Provision on Antenatal Care Use in Rural Zambia. PLOS ONE. 2012 Oct 4;7(10):e46475.

9. Nyambe S, Lungowe S, Choolwe J, Patrick M, Charles M. Factors associated with late antenatal care booking: population based observations from the 2007 Zambia demographic and health survey. PAMJ [Internet]. 2016 Oct 24 [cited 2022 Aug 12];25(109). Available from: https://www.panafrican-med-journal.com/content/article/25/109/full

10. Central Statistical Office/Zambia, Ministry of Health/Zambia, Tropical Disease Research Centre/Zambia, University of Zambia. Zambia Demographic and Health Survey 2007 [Internet]. Calverton, Maryland, USA: Central Statistical Office/Zambia and Macro International; 2009. Available from: http://dhsprogram.com/pubs/pdf/FR211/FR211.pdf

11. Zambia Statistics Agency - ZSA, Ministry of Health - MOH, University Teaching Hospital Virology Laboratory - UTH-VL, ICF. Zambia Demographic and Health Survey 2018 [Internet]. Lusaka, Zambia: ZSA, MOH, UTH-VL and ICF; 2020. Available from: https://www.dhsprogram.com/pubs/pdf/FR361/FR361.pdf

12. Central Statistical Office/Zambia, Ministry of Health/Zambia, University of Zambia Teaching Hospital Virology Laboratory, University of Zambia Department of Population Studies, Tropical Diseases Research Centre/Zambia, ICF International. Zambia Demographic and Health Survey 2013-14 [Internet]. Rockville, Maryland, USA: Central Statistical Office/Zambia, Ministry of Health/Zambia, and ICF International; 2015. Available from: http://dhsprogram.com/pubs/pdf/FR304/FR304.pdf

13. Ng’ambi WF, Collins JH, Colbourn T, Mangal T, Phillips A, Kachale F, et al. Socio-demographic factors associated with early antenatal care visits among pregnant women in Malawi: 2004–2016. PLOS ONE. 2022 Feb 8;17(2):e0263650.

14. Croft Trevor N., Marshall Aileen M. J., Allen Courtney K., et al. 2018. Guide to DHS Statistics. Rockville, Maryland, USA: ICF. Availible from: https://www.dhsprogram.com/pubs/pdf/DHSG1/Guide_to_DHS_Statistics_DHS-7_v2.pdf.

15. Friedman EM, Jang D. & Williams TV. (2002) Combined estimates from four quarterly survey data sets. Proceedings of the American Statistical Association Joint Statistical Meetings—Section on Survey Research Methods, pp.1064–1069. Alexandria, VA: American Statistical Association.

16. Ministry of Health. ANC GUIDELINES FOR A POSITIVE PREGNACY EXPERIENCE [Internet]. Lusaka, Zambia; 2022. Available from: https://www.afro.who.int/sites/default/files/2019-06/Draft%20ANC%20Guidelines%202018%20-%20Final%20Copy.pdf

17. Mlandu C, Matsena-Zingoni Z, Musenge E. Trends and determinants of late antenatal care initiation in three East African countries, 2007–2016: A population based cross-sectional analysis. PLOS Glob Public Health. 2022 Aug 15;2(8):e0000534.

18. WHO. WHO recommendation on antenatal care contact schedules [Internet]. WHO, Geneva, Switzerland; [cited 2022 Aug 18]. Available from: https://srhr.org/rhl/article/who-recommendation-on-antenatal-care-contact-schedules-2#:~:text=The%20focused%20ANC%20(FANC)%20model,between%2036%20and%2038%20weeks.

19. Ngoma-Hazemba A, Hamomba L, Silumbwe A, Munakampe MN, Soud F. Community Perspectives of a 3-Delays Model Intervention: A Qualitative Evaluation of Saving Mothers, Giving Life in Zambia. Glob Health Sci Pract. 2019 Mar 11;7(Supplement 1):S139.

20. CDC, USAID, PEPFAR, & SMGL. (2014). Saving Mothers, Giving Life: Maternal Mortality. Phase 1 Monitoring and Evaluation Report. Atlanta, GA: Centers for Disease Control and Prevention: US Dept of Health and Human Services.

21. Obse AG, Ataguba JE. Explaining socioeconomic disparities and gaps in the use of antenatal care services in 36 countries in sub-Saharan Africa. Health Policy Plan. 2021 Jun 3;36(5):651–61.

22. T. K. Sundari Ravindran & Veloshnee Govender (2020) Sexual and reproductive health services in universal health coverage: a review of recent evidence from lowand middle-income countries, Sexual and Reproductive Health Matters.

23. Gunn JKL, Asaolu IO, Center KE, Gibson SJ, Wightman P, Ezeanolue EE, et al. Antenatal care and uptake of HIV testing among pregnant women in sub-Saharan Africa: a cross-sectional study. J Int AIDS Soc. 2016;19(1):20605.

24. Ministry of Health. Zambia Consolidated Guidelines for Treatment and Prevention of HIV Infection [Internet]. MoH, Zambia; [cited 2022 Aug 18]. Available from: https://www.moh.gov.zm/wp-content/uploads/filebase/Zambia-Consolidated-Guidelines-for-Treatment-and-Prevention-of-HIV-Infection-2020.pdf

25. Consolidated guidelines on HIV prevention, testing, treatment, service delivery and monitoring: recommendations for a public health approach. Geneva: World Health Organization; 2021. Licence: CC BY-NC-SA 3.0 IGO.

26. Awopegba OE, Kalu A, Ahinkorah BO, Seidu AA, Ajayi AI. Prenatal care coverage and correlates of HIV testing in sub-Saharan Africa: Insight from demographic and health surveys of 16 countries. PLOS ONE. 2020 Nov 9;15(11):e0242001.

27. Staveteig S, Wang S, Head SK, Bradley SEK, Nybro E. Demographic patterns of HIV testing uptake in sub-Saharan Africa [Internet]. Calverton, Maryland, USA: ICF International; 2013. (DHS Comparative Reports No. 30). Available from: http://dhsprogram.com/pubs/pdf/CR30/CR30.pdf

28. UNAIDS. HIV testing and counselling [Internet]. UNAIDS; [cited 2022 Aug 18]. Available from: https://www.unaids.org/en/keywords/hiv-testing-and-counselling

